# Population vulnerability to COVID-19 in Europe: a burden of disease analysis

**DOI:** 10.1101/2020.04.29.20064279

**Authors:** Grant MA Wyper, Ricardo MA Assunção, Sarah Cuschieri, Brecht Devleeschauwer, Eilidh Fletcher, Juanita A Haagsma, Henk Hilderink, Jane Idavain, Tina Lesnik, Elena Von der Lippe, Marek Majdan, Milena S Milicevic, Elena Pallari, José L Peñalvo, Sara M Pires, Dietrich Plaß, João V Santos, Diane L Stockton, Sofie T Thomsen, Ian Grant

## Abstract

**Background:** Evidence has emerged showing that elderly people and those with pre-existing chronic health conditions may be at higher risk of developing severe health consequences from COVID-19. In Europe, this is of particular relevance with ageing populations living with non-communicable diseases, multi-morbidity and frailty. Published estimates of Years Lived with Disability (YLD) from the Global Burden of Disease (GBD) study help to characterise the extent of these effects. Our aim was to identify the countries across Europe that have populations at highest risk from COVID-19 by using estimates of population age structure and YLD for health conditions linked to severe illness from COVID-19.

**Methods:** Population and YLD estimates from GBD 2017 were extracted for 45 countries in Europe. YLD was restricted to a list of specific health conditions associated with being at risk of developing severe consequences from COVID-19 based on guidance from the United Kingdom Government. This guidance also identified individuals aged 70 years and above as being at higher risk of developing severe health consequences. Study outcomes were defined as: (i) proportion of population aged 70 years and above; and (ii) rate of YLD for COVID-19 for vulnerable health conditions across all ages. Bivariate groupings were established for each outcome and combined to establish overall population-level vulnerability.

**Results:** Countries with the highest proportions of elderly residents were Italy, Greece, Germany, Portugal and Finland. When assessments of population-level YLD rates for COVID-19 vulnerable health conditions were made the highest rates were observed for Bulgaria, Czech Republic, Croatia, Hungary and Bosnia and Herzegovina. A bivariate analysis indicated that the countries at high-risk across both measures of vulnerability were: Bulgaria; Portugal; Latvia; Lithuania; Greece; Germany; Estonia; and Sweden.

**Conclusion:** Routine estimates of population structures and non-fatal burden of disease measures can be usefully combined to create composite indicators of vulnerability for rapid assessments, in this case to severe health consequences from COVID-19. Countries with available results for sub-national regions within their country, or national burden of disease studies that also use sub-national levels for burden quantifications, should consider using non-fatal burden of disease estimates to estimate geographical vulnerability to COVID-19.

## Main paper

### Background

In burden of disease studies estimates of disability-adjusted life years (DALYs) are commonly used to assess the leading causes of burden amongst populations [1]. DALYs are composed of estimates of population health loss due to living with the consequences of morbidity and premature mortality. Years Lived with Disability (YLD) capture the morbidity (both the prevalence and severity of the disease) component of DALYs by estimating the number of years lost due to conditions diminishing the overall health status, and are a useful indicator to assess how impaired populations are due to living with the consequences of disease and injury [2].

Internationally, countries have reacted to the COVID-19 outbreak by introducing key public health non-pharmaceutical interventions (otherwise known as physical, or social, distancing) to protect vulnerable population groups [3]. Evidence has emerged to show that elderly people and those with pre-existing multi-morbid conditions may be at higher risk of developing severe health consequences from COVID-19 [4]. In Europe, 31% of the population are estimated to have a condition that is on the Government of the United Kingdom’s (UK) list of conditions at increased risk of severe health consequences from COVID-19 disease [5]. There is currently a disparity of comparable information across countries to objectively assess country-level vulnerability to COVID-19. However, there is a wealth of data on population structure, health status and causes of health loss in countries, which can be obtained from the Global Burden of Disease (GBD) study [6]. These data can be used to approximate how vulnerable populations are, particularly by focusing on the population share of elderly residents and the YLD for health conditions that have been identified as potentially linked to severe illness from COVID-19. This is of particular relevance for European countries, as increases in lifespan have resulted in increasingly ageing populations living with effects of non-communicable diseases, multi-morbidity and frailty [7].

The aim of this study was to identify the countries across Europe that have populations at highest risk for severe disease progression after COVID-19 infection by using estimates of population structure and YLD for health conditions linked to severe illness from COVID-19. This study was carried out using data from GBD 2017 for the reference year 2017, considering two measures of vulnerability: (i) rate of elderly population; and (ii) rate of YLD for health conditions identified at risk of severe health consequences from COVID-19.

### Methods

#### Data

The GBD Results Tool [8] was used to extract Years Lived with Disability (YLD) estimates for both sexes, age-groups (all ages; 70 years and above; and 80 years and above) and GBD 2017 level 3 cause [9] for each country defined as residing in Central, Eastern and Western Europe (N=45 countries). Estimates were considered for the constituent nations of the United Kingdom (UK): England; Northern Ireland; Scotland; and Wales, rather than the UK as a whole. In this study, hereafter, the elderly population denotes the age-group 70 years and above.

Data were retained for specific causes based on guidance from the UK Government (as at 30^th^ March 2020) on those health conditions that indicated a risk of severe health consequences from COVID-19 [10]. Two groups were defined: individuals aged 70 years and above, and those under 70 years that have one or more pre-existing underlying health condition. The guidance provided by the UK Government is outlined in the Supplementary Appendix and the list of pre-existing conditions were mapped to the GBD 2017 cause list (Table 1).

**Table 1.** Mapping of UK Government guidance on pre-existing medical conditions at risk of severe illness from COVID-19 to the GBD 2017 cause list [10]

| Pre-existing health condition(s) | GBD mapped cause(s) |
| --- | --- |
| All health conditions | All-causes |
| Chronic respiratory diseases | Chronic obstructive pulmonary disease<br>Pneumoconiosis<br>Asthma<br>Interstitial lung disease and pulmonary sarcoidosis<br>Other chronic respiratory diseases |
| Chronic heart disease | Cardiovascular diseases (chronic and acute) |
| Chronic kidney disease | Chronic kidney disease |
| Chronic liver disease | Cirrhosis and other chronic liver diseases |
| Chronic neurological conditions | Alzheimer's disease and other dementias<br>Parkinson's disease<br>Epilepsy<br>Multiple sclerosis<br>Motor neuron disease<br>Other neurological disorders |
| Diabetes | Diabetes mellitus |
| Problems with spleen | Sickle cell disorders<br>Sickle cell trait |
| Cancer undergoing active chemotherapy or radiotherapy | All cancer types |
| Cancers of the blood or bone marrow |  |
| Seriously overweight | Covered in mapping for chronic respiratory diseases, cardiovascular diseases, chronic kidney disease, cirrhosis and chronic liver diseases, and diabetes |
| Severe chest conditions such as cystic fibrosis or severe asthma | Covered in mapping for chronic respiratory diseases |
| Organ transplant and remain on ongoing immunosuppression medication | Covered in mapping for chronic kidney disease |
| Severe disease of body systems | Covered in mapping from all pre-existing health conditions |
| Those who are pregnant | Not covered |

Some emerging evidence has considered obesity and hypertension as independent risk factors for severe health consequences from COVID-19 [11, 12]. However, we do not consider them separately in this study as the vast majority of disease outcomes associated with these risk factors are included in the mapping to the GBD cause list (Table 1). For hypertension, all disease outcomes linked to high systolic blood pressure risk factor are included (cardiovascular diseases and chronic kidney disease in Table 1). In addition, GBD include separate estimates for hypertensive disease and these are included within cardiovascular diseases. The disease outcomes associated with obesity are also all covered in the mapping to the GBD cause list with the exception of Gout. Gout accounted for only 0.2% (95% uncertainty interval: 0.15-0.25%) of total YLD in GBD European Region in 2017 [8].

A permalink to the GBD Results Tool [8] query that were used to generate the data used in this study are outlined in the Data Availability section. Additionally, data on the total 2017 resident populations and population aged 70 years and above for each country were sourced from the Global Health Data Exchange (GHDx) [13]. These population denominators that were used in the production of GBD 2017 estimates.

#### Analyses

Descriptive summaries were calculated for the proportion of elderly population, and YLD for COVID-19 vulnerable health conditions were described using crude rates per 100,000 population. The numerators for the population proportion calculations were based on elderly populations, whereas the YLD rate calculation numerators were based on population totals. Denominators were based on the all ages population data sourced from GHDx [13].

Each measure was divided into tertiles (three binned categories: low; mid; and high). These categories were calculated to determine three equal size groups of vulnerability. Bivariate groupings were established by considering overlapping of the measures and were depicted in a scatter plot to identify groups of countries, both in terms of the proportion of elderly population and the rate of YLD for conditions associated with worse COVID-19 prognosis. Spearman’s rank correlation coefficient (ρ) was used to describe the correlation between the percentage of elderly population and the rate of YLD for COVID-19 vulnerable health conditions.

### Results

#### Proportions of elderly population by country

The five countries with the highest proportions of elderly residents (aged 70 years and above) were: Italy (16.4%); Greece (16.2%); Germany (15.4%); Portugal (15.3%); and Finland (14.7%) (Table 2). Conversely, the countries with the lowest proportions of elderly population were Israel (7.7%); Moldova (8.2%); Russian Federation (8.8%); Macedonia (8.8%); and Albania (9.0%). The ratio of the country with the highest (Italy) and lowest (Israel) proportion of elderly residents was 2.14, indicating over a two-fold difference between the countries.

**Table 2.**
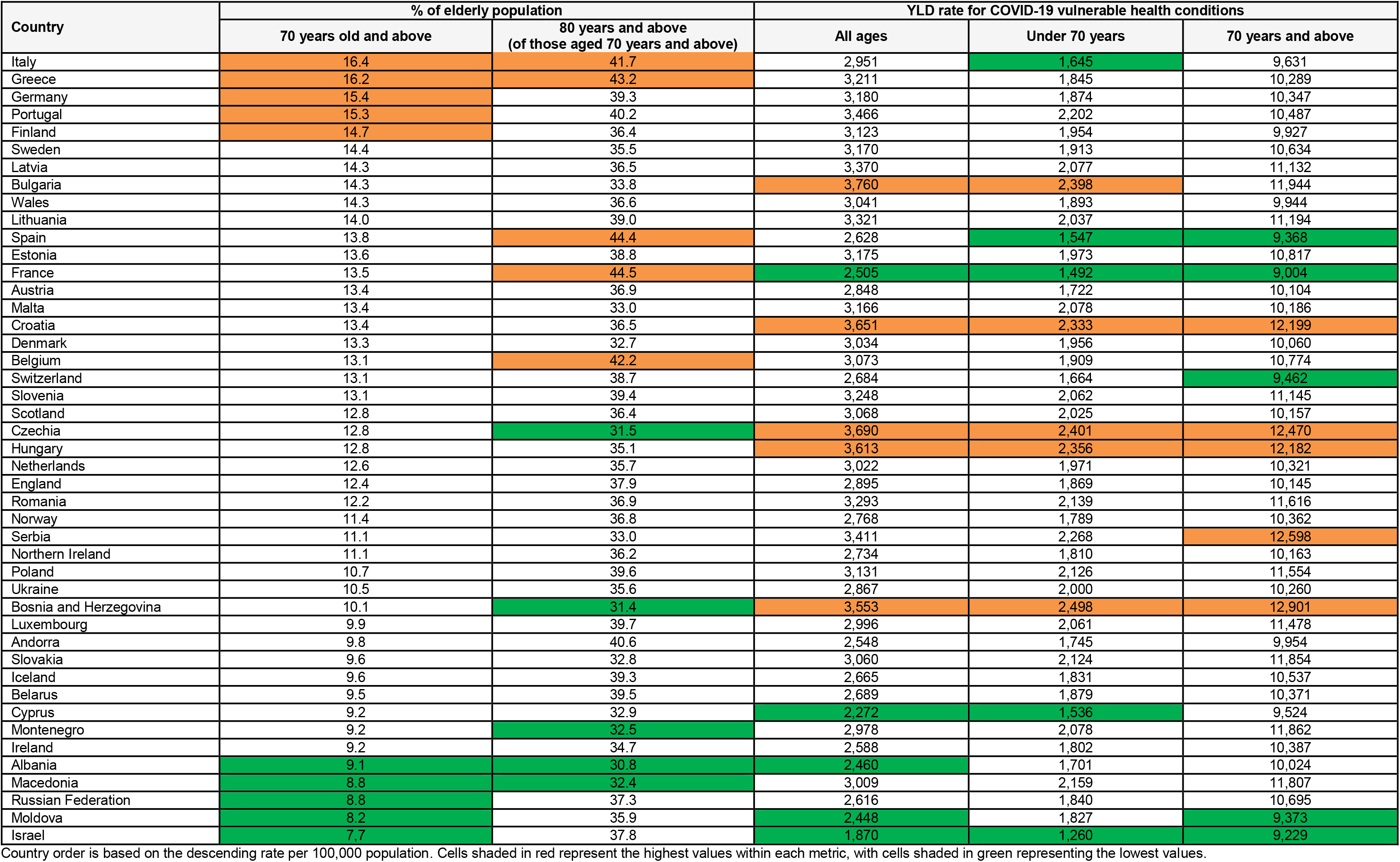
Summary of percentage of elderly population and YLD rates for COVID-19 vulnerable health conditions, by country, 2017

| Country | % of elderly population |  | YLD rate for COVID-19 vulnerable health conditions |  |  |
| --- | --- | --- | --- | --- | --- |
|  | 70 years old and above | 80 years and above<br>(of those aged 70 years and above) | All ages | Under 70 years | 70 years and above |
| Italy | 16.4 | 41.7 | 2,951 | 1,645 | 9,631 |
| Greece | 16.2 | 43.2 | 3,211 | 1,845 | 10,289 |
| Germany | 15.4 | 39.3 | 3,180 | 1,874 | 10,347 |
| Portugal | 15.3 | 40.2 | 3,466 | 2,202 | 10,487 |
| Finland | 14.7 | 36.4 | 3,123 | 1,954 | 9,927 |
| Sweden | 14.4 | 35.5 | 3,170 | 1,913 | 10,634 |
| Latvia | 14.3 | 36.5 | 3,370 | 2,077 | 11,132 |
| Bulgaria | 14.3 | 33.8 | 3,760 | 2,398 | 11,944 |
| Wales | 14.3 | 36.6 | 3,041 | 1,893 | 9,944 |
| Lithuania | 14.0 | 39.0 | 3,321 | 2,037 | 11,194 |
| Spain | 13.8 | 44.4 | 2,628 | 1,547 | 9,368 |
| Estonia | 13.6 | 38.8 | 3,175 | 1,973 | 10,817 |
| France | 13.5 | 44.5 | 2,505 | 1,492 | 9,004 |
| Austria | 13.4 | 36.9 | 2,848 | 1,722 | 10,104 |
| Malta | 13.4 | 33.0 | 3,166 | 2,078 | 10,186 |
| Croatia | 13.4 | 36.5 | 3,651 | 2,333 | 12,199 |
| Denmark | 13.3 | 32.7 | 3,034 | 1,956 | 10,060 |
| Belgium | 13.1 | 42.2 | 3,073 | 1,909 | 10,774 |
| Switzerland | 13.1 | 38.7 | 2,684 | 1,664 | 9,462 |
| Slovenia | 13.1 | 39.4 | 3,248 | 2,062 | 11,145 |
| Scotland | 12.8 | 36.4 | 3,068 | 2,025 | 10,157 |
| Czechia | 12.8 | 31.5 | 3,690 | 2,401 | 12,470 |
| Hungary | 12.8 | 35.1 | 3,613 | 2,356 | 12,182 |
| Netherlands | 12.6 | 35.7 | 3,022 | 1,971 | 10,321 |
| England | 12.4 | 37.9 | 2,895 | 1,869 | 10,145 |
| Romania | 12.2 | 36.9 | 3,293 | 2,139 | 11,616 |
| Norway | 11.4 | 36.8 | 2,768 | 1,789 | 10,362 |
| Serbia | 11.1 | 33.0 | 3,411 | 2,268 | 12,598 |
| Northern Ireland | 11.1 | 36.2 | 2,734 | 1,810 | 10,163 |
| Poland | 10.7 | 39.6 | 3,131 | 2,126 | 11,554 |
| Ukraine | 10.5 | 35.6 | 2,867 | 2,000 | 10,260 |
| Bosnia and Herzegovina | 10.1 | 31.4 | 3,553 | 2,498 | 12,901 |
| Luxembourg | 9.9 | 39.7 | 2,996 | 2,061 | 11,478 |
| Andorra | 9.8 | 40.6 | 2,548 | 1,745 | 9,954 |
| Slovakia | 9.6 | 32.8 | 3,060 | 2,124 | 11,854 |
| Iceland | 9.6 | 39.3 | 2,665 | 1,831 | 10,537 |
| Belarus | 9.5 | 39.5 | 2,689 | 1,879 | 10,371 |
| Cyprus | 9.2 | 32.9 | 2,272 | 1,536 | 9,524 |
| Montenegro | 9.2 | 32.5 | 2,978 | 2,078 | 11,862 |
| Ireland | 9.2 | 34.7 | 2,588 | 1,802 | 10,387 |
| Albania | 9.1 | 30.8 | 2,460 | 1,701 | 10,024 |
| Macedonia | 8.8 | 32.4 | 3,009 | 2,159 | 11,807 |
| Russian Federation | 8.8 | 37.3 | 2,616 | 1,840 | 10,695 |
| Moldova | 8.2 | 35.9 | 2,448 | 1,827 | 9,373 |
| Israel | 7.7 | 37.8 | 1,870 | 1,260 | 9,229 |

Country order is based on the descending rate per 100,000 population. Cells shaded in red represent the highest values within each metric, with cells shaded in green representing the lowest values.

When looking at stratified differences within the elderly age-group, the five countries with the highest percentage of population aged 80 years and above were: France (44.5%); Spain (44.4%); Greece (43.2%); Belgium (42.2%) and Italy (41.7%). The five countries with the lowest percentage of population aged 80 years and above were: Albania (30.8%); Bosnia and Herzegovina (31.4%); Czechia (31.5%); Macedonia (32.4%); and Montenegro (32.5%). Between the country with the highest (France) percentage of population aged 80 years and above and lowest (Albania), there was an absolute difference of 13.7%.

#### Rate of YLD for COVID-19 vulnerable health conditions

When the rate of YLD for health conditions associated with higher COVID-19 vulnerability was assessed for all ages, the five countries with the highest rates per 100,000 population were: Bulgaria (3,760); Czechia (3,690); Croatia (3,651), Hungary (3,613); and Bosnia and Herzegovina (3,553) (Table 2). The five countries with the lowest rates were: Israel (1,870); Cyprus (2,272); Moldova (2,448); Albania (2,460); and France (2,505). There was a rate ratio of 2.01 between the country with the highest rate (Bulgaria) and the country with the lowest rate (Israel).

Insights into rates of YLD for health conditions indicating higher COVID-19 vulnerability for those under 70 years and elderly residents were that there were four countries that were common amongst the leading five countries in both age-groups. These countries were: Czechia, Croatia, Hungary and Bosnia and Herzegovina. Of the five countries with the lowest rates in the under 70 years and elderly age-groups, there were three countries that were common: Israel, France and Spain.

#### Summary of combined vulnerability

There was a moderate association (ρ=0.54) between the percentage of elderly population and the rate of YLD for COVID-19 vulnerable health conditions. A bivariate analysis indicated that the countries which had high proportions of elderly population and high rates of YLD for COVID-19 vulnerable health conditions were: Bulgaria; Portugal; Latvia; Lithuania; Greece; Germany; Estonia; and Sweden. Conversely, the countries with the lowest proportions of elderly population and lowest rates of YLD for COVID-19 vulnerable health conditions were: Israel; Cyprus; Moldova; Albania; Andorra; Ireland; Russian Federation; Iceland; and Belarus. Bosnia and Herzegovina had a high rate of YLD for COVID-19 vulnerable health conditions, but a relatively low proportion of elderly population. On the other hand, Spain, France and Austria all had high proportions of elderly population but a relatively low rate of YLD for COVID-19 vulnerable health conditions (Figure 1).

**Fig. 1.**
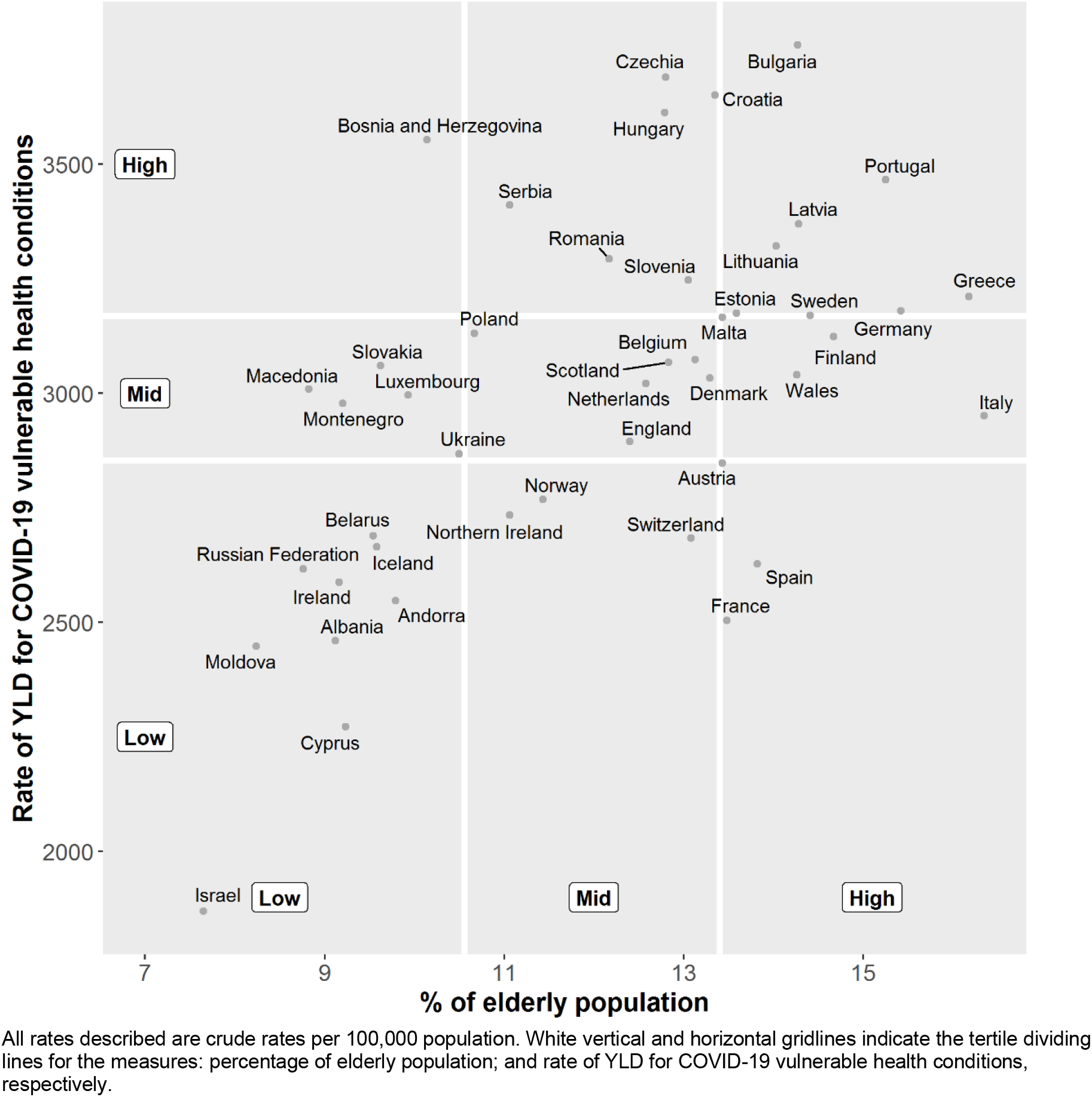
Scatter plot of percentage of elderly population versus rate of YLD for COVID-19 vulnerable health conditions for European countries

## Discussion

### Summary of findings

This study set out to establish which countries across Europe had populations that were most likely to be vulnerable to severe health consequences as a result of COVID-19 infection. This assessment was made using data on population age structure, and data on YLD for health conditions identified as increasing the risk of COVID-19 severity, the latter illustrating the extent to which populations are vulnerable through living with ill-health due to causes of disease.

Estimates of vulnerability to COVID-19 using elderly population share indicated that the countries with the highest proportions of elderly residents were Italy, Greece, Germany, Portugal and Finland. When assessments of population-level YLD rates for COVID-19 vulnerable health conditions were made the highest rates were observed for Bulgaria, Czechia, Croatia, Hungary and Bosnia and Herzegovina. Our bivariate analysis indicated that the countries which had high rates across both measures of vulnerability were: Bulgaria; Portugal; Latvia; Lithuania; Greece; Germany; Estonia; and Sweden.

Whilst these findings indicate population-level vulnerability due to health loss suffered, they do not take into account other important factors such as country and sub-national responses to the COVID-19 outbreak through public health non-pharmaceutical interventions. Neither do they take into account factors such as: population density, the capacity or ease of access to health and social care services and the disruption to existing services due to the COVID-19 crisis, all of which will have a significant impact on the extent to which vulnerable populations are adequately protected from harm. This may partly explain why countries identified in this analysis with high and low vulnerability to severe health consequences from COVID-19 do not always correspond with those countries in Europe with the highest and lowest case fatality ratios due to COVID-19 [14]. For example, within the Baltic states Latvia and Estonia have high vulnerability as measured on both indicators. However, Latvia responded to the crisis quickly by closing their borders and implementing restrictive measures much faster than Estonia, and case fatality rates are higher in Estonia [14–16]. This example highlights that a number of additional factors could contribute to differences between vulnerability and extent of adverse consequences, including: care identification and under-reporting, the speed at which countries introduced restrictive measures, and restrictions on air travel. The use of summary health indicator such as YLD to identify severe health consequences from COVID-19 infections should be regarded as just one of the elements that need to be taken into account in a complete risk assessment of vulnerability.

### Strengths and limitations

The study was carried out using estimates from GBD 2017, which is a widely used and well-established mechanism that has methodological consistency when producing estimates for individual countries [6]. The use of GBD 2017 is advantageous as estimates are publically accessible, which allows for the rapid assessments of the impact in response to public health emergency scenarios, such as the COVID-19 outbreak. Our findings are comparable on a like-for-like basis across countries. However, data sources that are fed into the modelling process for country-level estimates can vary based on location, therefore there is a risk that some of the differences which we observe may be attributed to the use, or omission, of high quality data sources [17]. We have opted not to include estimates of uncertainty in our estimates. Uncertainty intervals in the GBD study can often be wide, representing large degrees of uncertainty, so users of these results must bear in mind that these findings relate to the best available point-estimate. To retain consistency with estimates of YLD from GBD 2017, data on population size and structure was obtained from GHDx [13] which may differ from nationally produced estimates.

Previous research has suggested that the assumption of fixed severity distributions across countries may be unreasonable [18]. In our study of COVID-19 related vulnerable conditions, we did not include some of the leading causes of YLD, such as major depressive disorders and substance use disorders, which are thought to be the most likely to be affected by this assumption. Thus, our COVID-19 vulnerable conditions analysis may be less affected by this assumption [19]. Additionally, our study has assumed that the extent of vulnerability to COVID-19 can be determined by disability weights. For example, on average a greater weight would be given to those suffering from chronic obstructive pulmonary disease than to ischaemic heart disease [20]. This assumption may be problematic if the risk of COVID-19 associated with each health condition is not representative of relative differences in disability weight between causes. Also, particular combinations of disease may result in higher risks of consequences of COVID-19, while all combinations are in this approach assumed to have a similar effect.

We have used YLD as a proxy for the severity of the selected vulnerable health conditions as YLD includes a weighting of the severity of diseases stages i.e. a weighted prevalence. We have chosen to explore the aim of the YLD summary measure to combine all conditions, rather than examine the impact of individual causes. We acknowledge that using disease prevalence data from GHDx could add further insight into quantifying the disease specific implications of severe health consequences from COVID-19. However, since prevalence gives equal weighting to each condition, we did not consider prevalence as useful for summary analyses as YLD which allows a weighted sum of prevalence of different diseases. Further analysis has previously been carried out elsewhere to explore using prevalence to quantify the risk for severe health consequences from COVID-19 infection to enhance assessment of a health systems vulnerability to COVID-19 [5].

### Summary

Our findings have highlighted that routine data on population structure might be usefully extended by using estimates of YLD to consider how populations are impaired by living with the consequences of ill-health due to causes of disease and injury. Countries with available estimates for sub-national regions within their country, or national burden of disease studies that also estimate at sub-national levels should consider using non-fatal burden of disease estimates to estimate geographical vulnerability to COVID-19.

## Data Availability

The datasets used in this research study are all publically available.

http://ghdx.healthdata.org/gbd-results-tool?params=gbd-api-2017-permalink/376d9a9ad8401f49f104650fab0b9305

## List of abbreviations

COVID-19: Coronavirus Disease 2019
DALYs: Disability-Adjusted Life Years
GBD: Global Burden of Disease
GHDx: Global Health Data Exchange
UK: United Kingdom
YLD: Years Lived with Disability

## Declarations

### Ethics approval and consent to participate

Not applicable.

### Consent for publication

Not applicable.

### Availability of data and materials

The datasets used in this research study are all publically available. The permalink to data query used to obtain estimates of YLD is: http://ghdx.healthdata.org/gbd-results-tool?params=gbd-api-2017-permalink/376d9a9ad8401f49f104650fab0b9305

### Authors’ contributions

GW and IG generated the initial idea for the study. GW carried out all analyses and visualisation of the results. GW drafted the manuscript with assistance from IG. IG coordinated and made edits to the manuscript based on responses from co-authors with assistance from GW. All other authors provided critical input into the interpretation of the results, revisions to the manuscript and approved the final draft.

## Acknowledgements

We would like to acknowledge the GBD 2017 study investigators and collaborators, as without this study and their resulting estimates of population and YLD our study would not have been possible. Additionally, we would like to thank COST action CA18218 (European Burden of Disease Network) for bringing together researchers working across Europe on burden of disease activities, which has made this collaborative research possible.

## Supplementary Appendix

Guidance on social distancing for everyone in the United Kingdom

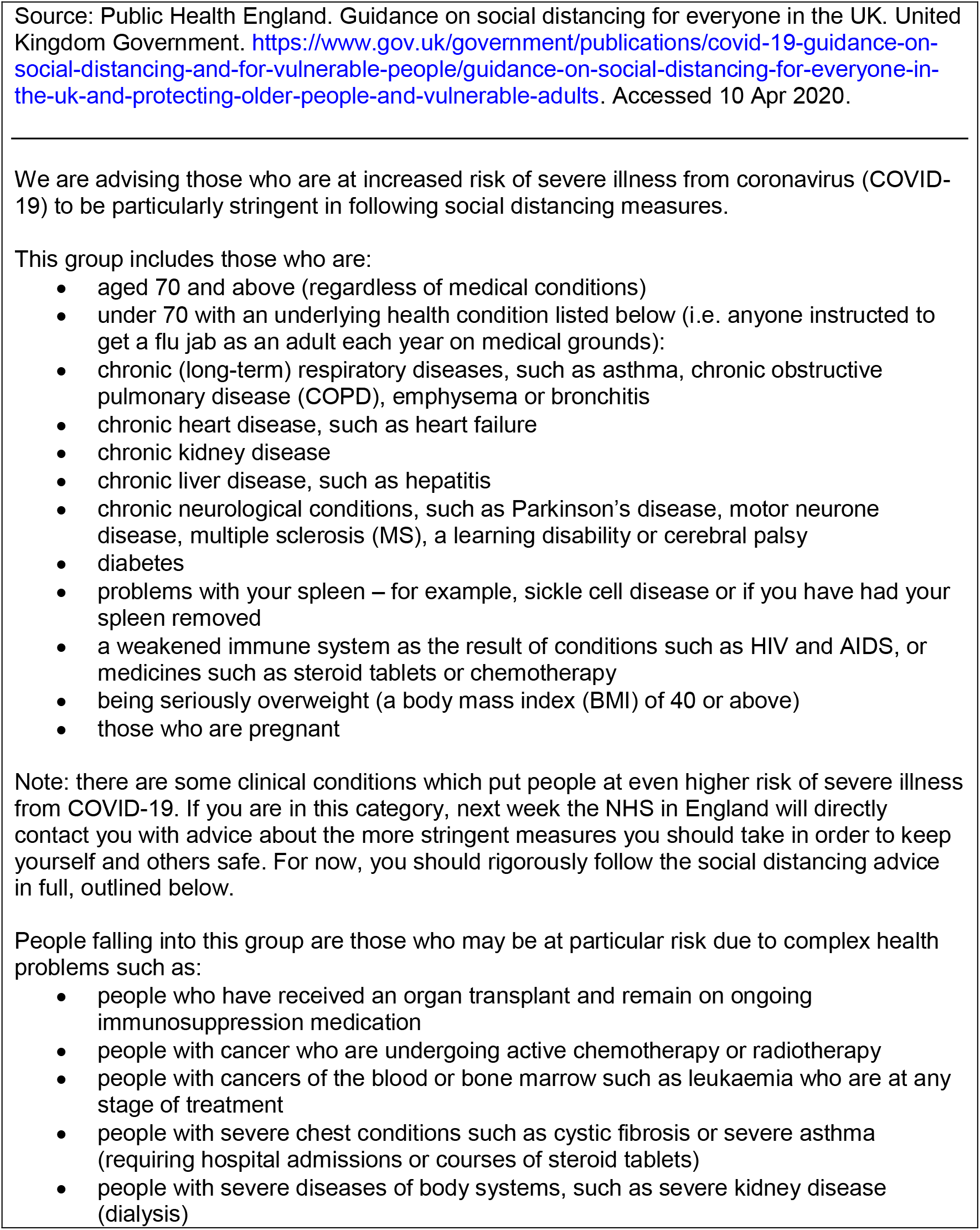

## References

1. Ware M, Mabe M. The STM Report An Overview of Scientific and Scholarly Journal Publishing Mark Ware Consulting International Association of Scientifi Fic, Technical and Medical Publishers.; 2015. www.markwareconsulting.com.

2. Hirsch JE. An index to quantify an individual’s scientific research output. Proc Natl Acad Sci. 2005. doi:10.1073/pnas.0507655102

3. Moed HF, Burger WJM, Frankfort JG, Van Raan AFJ. The application of bibliometric indicators: Important field- and time-dependent factors to be considered. Scientometrics. 1985. doi:10.1007/BF02016935

4. Lee JD, Cassano-Pinché A, Vicente KJ. Bibliometric analysis of Human Factors (1970–2000): A quantitative description of scientific impact. Hum Factors. 2005;47(4):753–766. doi:10.1518/001872005775570970

5. Davis PM. Open access, readership, citations: a randomized controlled trial of scientific journal publishing. FASEB J. 2011. doi:10.1096/fj.11_183988

6. Davis PM, Lewenstein BV, Simon DH, Booth JG, Connolly MJL. Open access publishing, article downloads, and citations: randomised controlled trial. BMJ. 2008;337:a568. doi:10.1136/bmj.a568

7. Antoniou GA, Antoniou SA, Georgakarakos EI, Sfyroeras GS, Georgiadis GS. Bibliometric analysis of factors predicting increased citations in the vascular and endovascular literature. Ann Vasc Surg. 2015;29(2):286–292. doi:10.1016/j.avsg.2014.09.017

8. Shekhani HN, Shariff S, Bhulani N, Khosa F, Hanna TN. Bibliometric Analysis of Manuscript Characteristics That Influence Citations: A Comparison of Six Major Radiology Journals. AJR Am J Roentgenol. 2017;209(6):1191–1196. doi:10.2214/AJR.17.18077

9. Aksnes DW. Characteristics of highly cited papers. Res Eval. 2003;12(3):159–170. doi:10.3152/147154403781776645

10. Falagas ME, Zarkali A, Karageorgopoulos DE, Bardakas V, Mavros MN. The Impact of Article Length on the Number of Future Citations: A Bibliometric Analysis of General Medicine Journals. PLoS One. 2013. doi:10.1371/journal.pone.0049476

11. Hafeez DM, Jalal S, Khosa F. Bibliometric analysis of manuscript characteristics that influence citations: A comparison of six major psychiatry journals. J Psychiatr Res. 2019. doi:10.1016/j.jpsychires.2018.07.010

12. Jacques TS, Sebire NJ. The impact of article titles on citation hits: an analysis of general and specialist medical journals. JRSM Short Rep. 2010;1(1):2. doi:10.1258/shorts.2009.100020

13. Paiva C, Lima J, Paiva B. Articles with short titles describing the results are cited more often. Clinics. 2012. doi:10.6061/clinics/2012(05)17

14. Habibzadeh F, Yadollahie M. Are Shorter Article Titles More Attractive for Citations? Cross-sectional Study of 22 Scientific Journals. Croat Med J. 2010. doi:10.3325/cmj.2010.51.165

15. Chokshi FH, Kang J, Kundu S, Castillo M. Bibliometric Analysis of Manuscript Title Characteristics Associated With Higher Citation Numbers: A Comparison of Three Major Radiology Journals, AJNR, AJR, and Radiology. Curr Probl Diagn Radiol. 45(6):356–360. doi:10.1067/j.cpradiol.2016.03.002

16. Nakayama T, Hirai N, Yamazaki S, Naito M. Adoption of structured abstracts by general medical journals and format for a structured abstract. J Med Libr Assoc. 2005.

17. Falagas ME, Pitsouni EI, Malietzis GA, Pappas G. Comparison of PubMed, Scopus, Web of Science, and Google Scholar: strengths and weaknesses. FASEB J. 2008;22(2):338–342. doi:10.1096/fj.07_9492LSF

18. Dunikowski LG, Freeman TR. Impact of family medicine research: Bibliometrics and beyond. Can Fam Physician. 2016;62(3):266–268. http://www.ncbi.nlm.nih.gov/pubmed/26975920. Accessed July 29, 2019.

